# Operationalization of the learned non-use phenomenon - A Delphi study

**DOI:** 10.1101/2020.03.18.20037374

**Authors:** Theresa Hirsch, Maria Barthel, Pauline Aarts, Yi-An Chen, Susanna Freivogel, Michelle J. Johnson, Theresa A. Jones, Marijtje L.A. Jongsma, Martina Maier, David Punt, Annette Sterr, Steven L. Wolf, Kirstin-Friederike Heise

**Affiliations:** University of Applied Sciences and Arts, Faculty of Social Work and Health, Hildesheim, Germany; University of Applied Sciences and Arts, Faculty of Natural Sciences and Technology, Goettingen, Germany; Sint Maartenskliniek, Department of Pediatric Rehabilitation, Nijmegen, The Netherlands; Georgia State University, Department of Occupational Therapy, Atlanta GA, USA; Department for Clinical Neurosciences and Preventive Medicine, Danube University Krems, Austria; University of Pennsylvania, Department of Physical Medicine and Rehabilitation, Philadelphia, PA, USA; Psychology Department and Neuroscience Institute, University of Texas at Austin, Austin, TX, USA; Behavioural Science Institute, Radboud University, Nijmegen, The Netherlands; Laboratory of Synthetic, Perceptive, Emotive and Cognitive Systems (SPECS), Institute for Bioengineering of Catalonia (IBEC), The Barcelona Institute of Science and Technology, Barcelona, Spain; School of Sport, Exercise & Rehabilitation Sciences, University of Birmingham, UK; School of Psychology, University of Surrey, Guildford, UK; Center for Postacute Neurorehabilitation, Berlin, Germany; Department of Rehabilitation Medicine, Emory University School of Medicine, Atlanta, GA, USA; Research Center for Movement Control and Neuroplasticity, Department of Movement Sciences, KU Leuven, Leuven, Belgium

**Author notes:** **Corresponding author** Kirstin-Friederike Heise, Research Center for Movement Control and Neuroplasticity, Department of Movement Sciences, Leuven, Belgium, Tervuursevest 101 box 1501, 3001 Leuven, Belgium. These authors contributed equally to the work (Theresa Hirsch, Maria Barthel).

**Keywords:** experience-dependent non-use, sensorimotor learning, rehabilitation, perceptual disorders, diagnosis

## Abstract

The discrepancy between residual functional capacity and reduced use of the contralesional hand, frequently observed after a brain lesion, has been termed Learned Non-Use (LNU) and is thought to depend on the interaction of neuronal mechanisms during recovery and learning-dependent mechanisms such as negative reinforcement. Despite the generally accepted existence of the LNU phenomenon among clinicians and researchers, no unequivocal and transdisciplinary definition exists to date. Furthermore, although therapeutic approaches are implemented in clinical practice to explicitly target LNU, no standardized diagnostic routine is described in the current literature.

Based on a structured group communication following the Delphi method among clinical and scientific experts in the field of LNU, knowledge from both, the work with patient populations and with animal models, was synthesized and integrated to reach consensus regarding a transdisciplinary definition of the LNU phenomenon. Furthermore, the mode and strategy of the diagnostic process, as well as the sources of information and outcome parameters relevant for the clinical decision making, were described with a wide range showing the current lack of a consistent universal diagnostic approach. Building on these results, the need for the development of a structured diagnostic procedure and its implementation into clinical practice is emphasized. Moreover, it exists a striking gap between the prevailing hypotheses regarding the mechanisms underlying the LNU phenomenon and the actual evidence. Therefore, basic research is needed to bridge between bedside and bench and eventually improve clinical decision making and further development of interventional strategies beyond the field of stroke rehabilitation.

## Introduction

Stroke is among the leading causes of long-term disability in adults and the worldwide population of stroke survivors is predicted to increase to 70 million within the next decade, clearly challenging national health care systems (Feigin et al., 2014). One of the most disabling and persisting consequences of stroke is the paresis of the upper extremity (hand/arm) (Jørgensen et al., 1995; Lai et al., 2002).

One frequent clinical observation is a decreased use of the contralesional hand in spite of latent functional ability and an increased functional reliance on the less-/non-impaired upper extremity over time (Sterr et al., 2002). This discrepancy between residual functional capabilities of the impaired limb and yet reduced usage in every-day life has been termed Learned Non-Use (LNU) and is assumed to be brought about by an interaction between post-lesion neural reorganization and mechanisms such as negative reinforcement (Taub, 1980; Allred et al., 2014). This conceptual framework goes back to the work of Charles Sherrington and colleagues who observed the reduced upper extremity use after surgical interruption of the sensory tracts (de-afferentation) in monkeys (Mott and Sherrington, 1894; Sherrington, 1931). Following up on this seminal work, Edward Taub and co-workers were the first to suggest learning psychological mechanisms to be involved in the development and maintenance of functional disuse of the deafferented forelimb (Taub, 1976b; Taub, 1976a). They subsequently transferred the LNU concept into the field of neurorehabilitation and developed an intensive training regimen (initially termed Forced Use Therapy) targeted at counteracting the non-use phenomenon and regaining the lost upper-extremity function [reviewed in (Taub et al., 2014)]. This work has inspired ample experimental and clinical work analyzing the effect of massed practice of the affected upper extremity in humans recovering from stroke (Corbetta et al., 2015). Despite the success of this therapeutic approach in the acute and most remarkably in the chronic stage, the definition of LNU has remained rather indistinct throughout the years. Moreover, the diagnostic verification of the actual presence of LNU has received little attention and is oftentimes left to anecdotal findings and subjective descriptions of the patient’s behavior.

In summary, no transdisciplinary (Stember, 1991; Baur and Blasius, 2014b) definition exists to date, which would allow a clear and unambiguous description of the LNU phenomenon. Furthermore, so far, no gold standard diagnostic tool exists allowing to identify the LNU phenomenon in differentiation from other alternative symptom complexes, which clearly obscures the evaluation of the effect of therapeutic interventions in clinical populations.

Therefore, the objective of the present work was two-fold: Firstly, to generate a comprehensive transdisciplinary definition of the LNU phenomenon; and secondly, to facilitate a synthesis of the available assessment strategies and required methodological components for the diagnosis of LNU in patient populations. We, therefore, chose the Delphi method as a structured group communication among clinical and scientific experts in the field of LNU, aiming at integrating the knowledge base resulting from work in healthy and clinical human populations as well as from work with animal models.

## Method

The Delphi technique is defined as a group communication process involving an interaction between the researcher and a group of identified experts on a specified topic, usually through a series of questions or questionnaires (Häder, 2009; Baur and Blasius, 2014a). The Delphi method was developed by Olaf Helmer and his colleagues at the Rand Corporation in the early 1950s (Helmer-Hirschberg, 1967) and has been used to gain a consensus regarding future trends and projections using a systematic process of information gathering.

### Selection of Participants as Experts

Experts for the present study are defined as being author/co-author of international peer-reviewed publication on the focus topic “learned nonuse” identified by a database (PubMed, EMBASE) search [combinations of key words: learned nonuse, experience dependent nonuse, disuse, compensation, stroke, brain lesion, upper extremity, upper limb, forelimb, human, stroke model]. The identified experts were contacted by email (see below) in English and German language.

We decided to offer the co-authorship of the expected publication of the planned Delphi study’s results for two main reasons: (1.) Following the example of published work in the field of rehabilitation [e.g. (Luedtke et al., 2016)], our reasoning was that offering a co-authorship represents a valid and appropriate acknowledgement of the participants’ time investment. (2.) Since the results of the Delphi study vitally depend on the experts’ motivation to share their expertise and opinion throughout several rounds of the Delphi process, we aimed at assuring constant adherence throughout the study as well as meaningful responses by offering a co-authorship. The direct involvement of the respondents’ motivation in the topic of concern and their high-level motivation of has been defined as a key element to the success and value of a Delphi study (Millar et al., 2007).

We regard the potential bias that this procedure might incorporate as limited and of minimal danger with respect to potential interference with the results of the study. Due to the Delphi study’s methodology that uses feedback of the answers to the participants after each round, we are confident that including the participants as co-authors has not diminish the value of the outcome and is strictly in line with the advantages of this methodical approach to capture and combine the knowledge and abilities of a diverse group of experts with the “democratic virtues of transparency and openness to criticism” as initially proposed for the Delphi method (Millar et al., 2007).

Experts identified according to the above-mentioned criteria were contacted via email two weeks before the planned start of the structured group communication process and asked to express their interest in participating in the Delphi study by replying to the email, i.e. active informed consent. Concise information on the purpose, the procedures of the study, as well as data protection was given in this first email to enable their general decision for or against participation in the Delphi study. All recipients of the initial email were informed that it would be possible for the research personnel directly involved in the data acquisition and analysis to map the answers to the individual participants. Automatic email responses were not considered and no response to the initial email was taken as withheld consent. After active response to the initial email, experts were registered as participants in the Delphi study and received the link to the online survey. From initially 41 identified international experts, 18 actively consented to participate in at least one round of questions. The final sample represented clinicians and scientists in the area of applied clinical research and basic sciences working with human and animal data in England, France, Germany, Netherlands, Spain, Switzerland, and the United States of America.

During the group communication process, the participants of the study remained anonymous towards each other. No personal data except the online available information on the experts (personal webpage, institutional webpage) was acquired. Since a co-authorship was offered to all experts who participated in the group communication, personal information (name and affiliations) is revealed of those who agreed to the co-authorship.

### Procedure

Based an initial literature search (03-04/2017) to identify experts described above and a second search (07/2017), we screened the retrieved literature to gather information about (1) existing descriptions of and (2) procedures to identify the LNU phenomenon. The resultant information was used to deduce a system of main categories, which served as the basis to formulate the questions for the first round of the group communication. The group communication was performed with an established online survey tool (SurveyMonkey Inc, San Mateo, California, USA, www.surveymonkey.com) that suffices common data protection regulations.

#### Textbox 1

**Initial Questions of first Delphi round**

1. *What is your definition of the Learned Non-Use phenomenon? Please describe in detail the Learned Non-Use phenomenon based on your expert knowledge*.
2. *Thinking about the optimal process for diagnosing Learned Non-Use of the hand in patients with a lesion of the central nervous system – what should it look like? Based on your expertise, please describe an optimal process of diagnosing Learned Non-Use (components, chronological processes, professional categories etc.)*.

Starting from two initial questions (textbox 1), the group communication followed an iterative process in which the responses of each question round were fed back to the study participants with the two main goals (1) to reach a consensus with respect the definition of the phenomenon and (2) to synthesize the components of the diagnostic process. The phase of the structured group communication comprised of three rounds of questions and responses and took place between 05/2017 and 09/2017 (overview of the full process of the Delphi study is given in figure 1).

**Figure 1.**
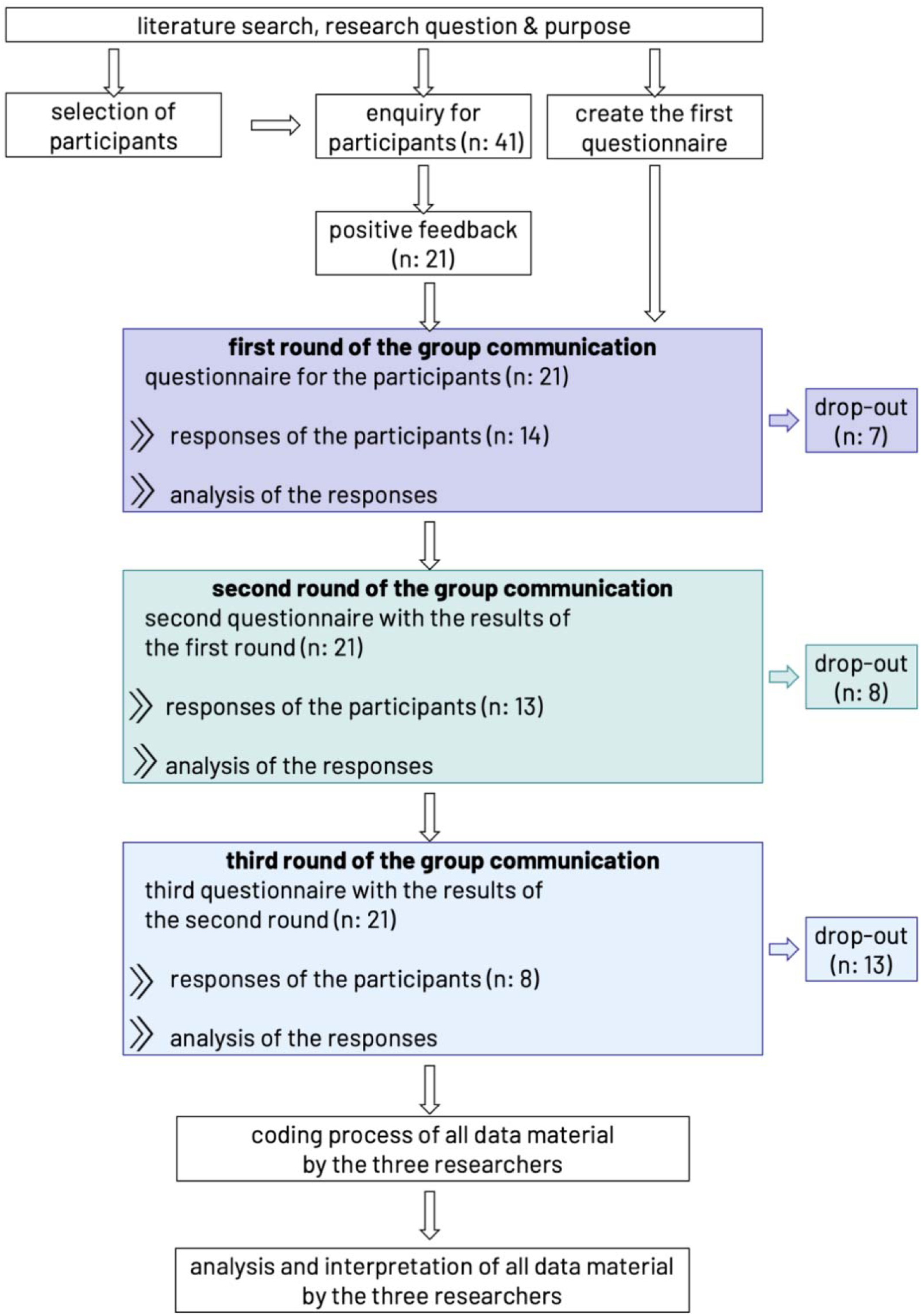
Flowchart of methodological approach of data acquisition and analysis and attrition of participating experts throughout the Delphi study.

### Data analysis

The data material gathered in the three rounds of open questions was subjected to a Qualitative Text Analysis following Kuckartz (Kuckartz, 2018), which consists of a category-based analysis approach of the text material. The main goal of the structured qualitative content analysis is defined as developing main and subcategories, which structure the content of the data and reduce the material allowing for consensus and synthesis. We chose this method because both, the research question and the goal require an exploratory approach in the data collection and analysis rather than the verification of an a priori hypothesis. Hence, the Qualitative Content Analysis allows a stepwise development of the central topics of the subject area, which is optimally suited for the systematic integration of deductively and inductively generated new knowledge and opinions under methodologically well-controlled conditions.

Starting from the a priori deductively developed system of main categories (see above), additional main and subcategories were inductively developed based on the data material. In the present case, the data material consisted of English and German responses, which were left in their original language in this first analysis phase in order to retain the original source information as closely as possible. This initiating analysis of the data material comprised of a first identification of important statements and the collection of main ideas with the overall objective to summarize the information. This phase was carried out by TH for all three Delphi rounds in communication with KFH and MB with the goal to extract the main information, to generate the feedback to the participants and to formulate the follow-up questions for the subsequent Delphi rounds. In a second phase, the main categories, which were a priori deductively developed from the available body of literature (see above), were complemented by additional main categories developed inductively from the data material after the third Delphi round. We followed an open coding strategy of newly developed topics. This step was carried out on approximately 10% of the data material in order to test the specific applicability of the existing categories for the present data material by MB and KFH independently from each other. Consensus regarding the main categories was achieved in a communicative process involving all three researchers (TH, MB, and KFH). In a third phase, the coding process of the complete data material was accomplished. Independently from each other, all three researchers (TH, MB, and KFH) coded all available data material focusing on defined sub-topics, which were related to a sub-set of research questions from all three Delphi rounds. Every single section of the available data material was admitted to coding and received single or multiple categories defined in phase 2. After this step, another communicative step among all three researchers (TH, MB, and KFH) yielded consensus regarding the coding of the respective others’ data material. As a result of this step, the category system was further specified, and the coding was re-evaluated. The next phase yielded a summary of the coded material and a further inductive definition of subcategories. The final phases comprised a second coding process of the whole data material geared at achieving a further differentiated category system. As an intermediate step, a systematic case summary was produced employing a framework analysis, which resulted in a theme matrix. By permuting the data within the category system, we aimed at condensing the data material and achieve a pointed description of aspects relevant to our research question. Finally, the analysis and interpretation of the data, including the visualization of results, constituted the last phase. For publication purposes, responses in German language were translated into English and translations were validated by all three researchers (TH, MB, and KFH). Despite the clear differentiation of the seven phases in this overview, it is necessary to point out that the individual phases were interleaved and partially repeated throughout the process of data analysis until consensus among the three researchers was achieved. We furthermore included an additional step involving the expert participants of the Delphi study to comment on the results and their final interpretation, which received recognition in the discussion section.

### Ethics Statement

The protocol and all procedures of this study complied with the ethical requirements as approved by the Social and Societal Ethics Committee of the KU Leuven (SMEC protocol number G-2017 06 841).

## Results

The questions formulated based on the initial literature search as well as the questions derived from the subsequent Delphi rounds are given in supplemental online material. As depicted in figure 1, the synthesis of the responses from one round was integrated into the questions of the subsequent round, and all available data material was analyzed in its entirety within each analysis step throughout the process. Therefore, the results are presented here in the permutated order of deductively and inductively developed main and subcategories and are hence generally free from the chronological order of data acquisition. Citations of original responses are presented in English, i.e., German responses are presented in their translated version without explicitly stating as such, and are referenced here with the internal identification scheme to keep the anonymity of participants.

### Definition LNU

The three rounds of questions brought about a wide range of supposedly defining elements of the LNU phenomenon. In this context, two main hypotheses related to the underlying mechanisms were mentioned. On the one hand, this consisted of the initial neuronal injury and the resulting structural impairment and reduction of sensorimotor functioning. On the other hand, experience-dependent processes secondary to the initial lesion were outlined to explain the observed change in behavior over time. In particular negative experiences, such as “*unsuccessful movements*” or “*unsuccessful attempts to move*” of the contralesional extremity were mentioned in this context by several participants.

All participants agreed on the main characteristic of the discrepancy between the actual spontaneous use and the residual function as the indicator for the conditioned non-use behavior.

Based on these elements, a first draft definition was fed back to the participants and a consensus regarding the definition was reached in the third round, suggested by one expert in its final version:

> *Learned non-use is the discrepancy between a retained motor capacity, which can be retrieved when requested, and the spontaneous use of this motor capacity in daily life, a discrepancy that develops due to experience in inept or non-productive perceived attempts to use that capacity*. [C6.r2]

A recurring theme, which was controversially discussed among the participants, was the aspect of compensatory use of the ipsilesional side as an inherent characteristic for the LNU phenomenon. On the one hand, compensation was discussed as an inevitable and logic step in the course of the development of new movement strategies and hence as a key marker for the identification of the LNU phenomenon.

> *„This phenomenon develops as the client attempts to use the involved extremity and is unsuccessful. He or she immediately is confronted with the choice to be dependent or unsuccessful when attempting to engage in functional activities or be creative and develop an alternative strategy for being successful in activities.”* [A5.r1]

The compensatory behavior was, on the other hand, hypothesized as a potential factor, which might additionally contribute to the manifestation of the non-use behavior.

> *“The person affected tries to move the paretic extremity. Hereupon negative feedback reaches the affected cortical area. Already after a short period of time and a couple of unsuccessful attempts to move, a ‘fading out’ of this cortical area happens. […] Instead the cortex switches to compensatory measures. A way to achieve the goal (e.g. reach to grasp objects) needs to be found. The most obvious solution is to use the other, the healthy […] extremity. This leads to positive feedback because our brain is functioning in a result-oriented way.”* [A3.r1]

However, compensatory behavior was also discussed as a strategy that is beneficial for regaining independence in daily life activities.

> *“[…] one must understand that many health care professionals immediately teach the client to be more ‘independent’ by showing them how to compensate with the uninvolved upper extremity in order to become self-sufficient. One can therefore regard learned non-use as learning an alternative strategy for the same activity. It can be seen as creative and a coping strategy. It is clear also that exactly these strategies are the reason that clients learn not to use the involved upper extremity even when the upper extremity and hand become more dynamic.”* [A5.r1]

Considering these additional aspects, the final definition was amended as follows:

Learned non-use is the discrepancy between a retained motor capacity, which can be retrieved when requested, and the spontaneous use of this motor capacity in daily life, a discrepancy that develops due to experience in inept or non-productive perceived attempts to use that capacity. It is accompanied by compensatory motor behavior identifiable in other body parts.

### Critical factors for the development of the phenomenon

It became clear from the answers related to the defining characteristics outlined before, that different assumptions prevail with respect to the factors, which are considered critical for the development of the LNU phenomenon. While the neuronal lesion overall was considered the precondition for the development, the participants’ answers showed less agreement regarding the influence of lesion location and size, severity of the initial impairment, or recovery time-course. The reorganization pattern following the lesion and in particular the degree of bilateral reorganization was assumed to determine and modify the development of non-use of the contralesional hand.

> “*In our opinion, it seems important to consider whether central lesions also affect the somatosensory system. Whether or not tactile / somatosensory / proprioceptive information processing is also affected might play a major part with respect to whether or not learned non-use occurs*.” [C1.r2]
>
> “*Severity of impairment”* [C6.r2]
>
> *„Extent of bilateral reorganization needed. […] bilaterality of the stroke - % involvement of the less affected side”* [C9.r2]
>
> “*the functional deficits following a stroke are strongly influenced by the lesion location and size; there are however patients with comparable lesion who show marked differences in their functional recovery*” [A4.r1]

Moreover, personal and contextual factors were mentioned with overall agreement as defining factors or effect modulators in the development of the non-use. With respect to personal factors, the individual’s psychological and cognitive situation was considered of particular relevance.

> „*motivation and patience of the patient*” [A2.r 2]
>
> “*psychological aspects associated with having a visible disability*” [C5.r2]
>
> *“Cognitive deficits or a form of dementia following in the context of the cerebro-vascular lesion render the development of the learned-nonuse more likely because those people affected have oftentimes not the abilities to process and retrieve the relevant background information.”* [A3.r2]

Likewise, premorbid factors as for example whether the contralesional hand was the dominant or non-dominant side were considered relevant individual factors contributing to the development of compensatory strategies and hence non-use behavior.

The self-efficacy in daily life and the individual’s situation of social support was mentioned as relevant contextual factors that could impact on the actual use of the contralesional extremity both, in a positive or negative way.

> „*informative consultation by the therapist*” [A2.r 2]
>
> “*informed family members*” [A3.r2]
>
> „*Social support*” [B3.r2]
>
> „*rehab settings focusing on ‘quick’ wins and often focus on compensatory strategies*” [C5.r2]

### LNU Diagnostic procedure

While it became clear in three rounds of questions that no generally agreed or standard procedure to diagnose the LNU phenomenon exists to date, the basic features of the optimal diagnostic process were consistently described in all three rounds. Additionally, the need for objectively quantifiable outcome parameters was stressed throughout the whole phase of data acquisition.

#### Process/approach

One significant criterion widely agreed upon was the multidisciplinary team, including all health professionals in contact with the patient, to be involved in the diagnostic process. Furthermore, the majority of the participants stressed the requirement of a longitudinal diagnostic time window spanning the subacute to the chronic phase after the initial lesion. Therefore, acute and rehabilitation clinical, as well as the home environment, were mentioned as potential settings.

The mode of diagnostic procedures was described somewhat multifaceted involving the enumeration of various existing standardized tests for sensorimotor functioning, observation of specific activities, interviews of patients and relatives, and the use of wearables and motion capture technology. Discussed were also the degree of continuity and sampling rate of the data to be acquired. The aspect of comparison of the integration of the contralesional hand into a defined task in different situations was discussed with two main settings, the spontaneous use and the forced or use upon request or instruction, to be compared.

> *“The optimal process for diagnosing Learned Non-Use would be to compare the spontaneous use of the hand in daily life activities (measured objectively) with the ability of the paretic hand to perform the same tasks when forced to do so (measured objectively). Learned Non-Use would be the difference between the objective score obtained in the laboratory when forced to use the paretic hand and the objective score obtained in spontaneous condition (daily life activities) with either hand paretic or non-paretic.”* [B1.r1] *“Observing what a patient does automatically vs. what they do when cued (forced) provides reasonable diagnostic value at the bedside”* [C2.r1] *“…*
>
> *learned non-use can be identified when: (1) a patient fails to make any spontaneous effort to use a limb in purposeful movement; or (2) efforts to activate the limb by immobilizing the contralateral limb (i.e*., *having the patient sit on it) and engaging subconscious activity (such as gesturing while talking) fails to evoke any spontaneous movement.”* [C8.r1]

In some descriptions of the diagnostic test strategies, typical therapeutic elements of the CIMT approach (sequencing, grading) were clearly discernable.

> “*In his performance upon request […], the patient shows us his automatic action and movement pattern. This is what is necessary to identify the learned non-use phenomenon. The patient should perform various bimanual tasks, graded from gross motor to sequencing of fine motor task. The therapist works hands-off in order to recognize the movement pattern in an unveiled fashion. […] The therapist observes if and how the affected hand is used or if the affected hand is generally neglected and only the healthy hand is worked with.”* [A3.r1]

In this context, the strategy of contrasting the spontaneous daily life situation with the explicit test situation was repeatedly discussed. Whether or not the patient should be aware of the test situation and the potential of implicit measurement methods were additionally raised as relevant aspects.

> *“We believe that the diagnosis of learned non-use can only be done through implicit measurement methods or tools that give an unbiased/ no subjective view on the patient’s status. The advancement of technology has helped us in this regard since it allows us to continuously obtain data with a much higher resolution and no human intervention. Specifically, we believe that wearables could be the ideal diagnostic tool for measuring learned non-use. Wearables allow measuring the level learned non-use at the patients’ home when performing everyday activities.”* [B2.r1]
>
> *“The patient will not be informed about the exact goal of the analysis (recognizing the learned non-use) until after the examination because this new automatic behaviour does not equally well appear in a test situation as it does in a daily life situation.”* [A3.r1]

In contradiction to present clinical practice, some of the participants sharply criticized interview-based evaluation methods relying on self-reported and memorized events. This group of participants was instead favoring prospective evaluation methods yielding unbiased information that does not rely on subjective evaluation criteria. The patient’s naivety towards the goal of the test and the avoidance of manual interference of the evaluator were also mentioned as necessary for objectifiable test results.

#### Sources of information

Different sources of information were detailed by the participants reflecting the variety of strategies currently in use. While spontaneous use and dis-/non-use of the contralesional hand in daily life were among the most often mentioned points, the patient’s subjective perspective concerning his/her movement quantity and quality received less frequent mentioning. The most compound approach was delineated as a combination of the aforementioned, i.e., data supporting both, the quantification of the nonuse as well as the retained functional capacity.

> *“(#1) data supporting the presence of non-use of some capacity and (#2) data supporting that this non-use followed non-rewarding attempts to use that capacity. In the absence of continuous monitoring, #2 would presumably have to rely on retrospective self-report, with its high potential to be faulty. One could instead (#3) establish there is good remaining capacity to use the non-used capacity, but this would leave open whether this non-use is learned. A reasonable solution might be to combine tests of #1, 2 & 3.”* [C6.r1]

The target dimensions, which the participants focus on in the diagnostic process, can be differentiated into body functions, activities, and daily use. Qualitative and quantitative measures were explained to be mutually considered for the evaluation of the respective dimensions. Among the dimensions stated were those directly related to the upper extremity (hand/arm muscle strength, movement parameters, movement attempts) but also those related to gross motor function, overall constitution, and the cognitive domain (learning, attention).

The direct relevance of the dimensions listed in the answers to the concept of LNU was not always explained and seemed generic to the evaluation of sensorimotor function.

#### Differential diagnosis

In order to differentiate the existence of the LNU phenomenon and exclude potential other symptoms, the participants mentioned established neurological, motor-functional, and neuropsychological tests. The leading indicator for the existence of the LNU phenomenon was considered the mainly behavioral phenomenon evidenced by the supposedly apparent discrepancy between spontaneous and forced use of the affected hand. Importantly, there was disagreement on whether the LNU phenomenon is at all differentiable from other syndromes such as motor neglect.

> *“Learned non-use is not a diagnostically related term nor does it exist within CPT codes or other classification systems for which reimbursement for services is provided.”* [C8.r1]
>
> “*I am not sure they can be* [differentiated]. *Motor neglect and learned nonuse share the same working definition.”* [C2.r2]
>
> “*I’m not sure that it can be entirely disentangled from motor neglect. Neglect is associated with non-use and CI therapy improves symptoms of hemispatial neglect. Apraxia is a deficit of motor planning and is an entirely different phenomenon, though it is possible that apraxia could increase risk for nonuse because it makes movement more effortful*.” [C3.r2]

#### Evidencing the experience-dependence of LNU symptoms

Depending on the prevailing assumptions about the underlying mechanisms of the LNU phenomenon, sensorimotor, mental, behavioral, and perceptive components were added by the participants to their description of the target dimensions. When investigating the participants’ assumptions regarding indications for the phenomenon’s origin in an experience- or learning-dependent process, it became apparent that no new information was added.

Instead, it appeared as if the theory of the contribution of learning mechanisms to the phenomenon’s development was hardly questioned in itself.

> “*One key element of learning is that short bouts of training will affect the outcomes. That is, if it is learned it can also be ‘unlearned’. So, a short learning study, in which aspects of the phenomenon are trained may inform on the ‘hard-wiredness’ of the phenomenon.”* [C4.r2]

Consequently, the description of outcome parameters targeting this assumed learning process remained vague and was only outlined very superficially.

> “*I think that the only way to get at this would be to track use attempts over time using wearable sensors or video recordings*.” [C3.r3]
>
> “*Kinematics difference between limb usage in coupled versus uncoupled bilateral symmetric tasks.”* [C9.r3]
>
> “[…] *ability to use a limb when explicitly needed but the lack of the spontaneous use of that limb*.” [C1.r3]

The explicit question tailored to explore this aspect further (see supplemental material round 3 question 1) did not yield sufficiently new information aside from the potential quantification of the discrepancy of the spontaneous versus the forced use over time as a potential readout of experience-dependent change over time, which was mentioned already in preceding question rounds.

> “*I think that the only way to get at this would be to track use attempts over time using wearable sensors or video recordings. If non-use is learning related, there should be a period of time in which use is greater post-stroke, followed by limited success, and then decreased attempts thereafter.”* [C6.r3]

Nonetheless, the lack of evidence for the experience-dependence of the non-use behavior in patients received a critical evaluation, and the necessity to build a proof for this fundamental assumption was strongly emphasized.

> *“There have been strong studies in rats and non-human primates that have characterized the learning process involved in learned non-use in the early period after CNS injuries. These are not paralleled by similar clinical studies. Such studies are desperately needed, both to firmly establish the degree to which learned non-use contributes to disuse and to start to reveal specific characteristics that could be used for differential diagnosis.”* [C6.r2]

Due to the saturation of the information acquired in response to this question, no further question round was initiated.

## Discussion

In three rounds of the present Delphi study, consensus could be reached on a transdisciplinary definition characterizing the learned non-use phenomenon. While the key elements defining the actual gap between spontaneous use and functional capacity of the contralesional extremity as well as the experience-dependence of this phenomenon were uncontentious among the participants, the role of the ipsilesional side was discussed controversially. In the original work building up to formulating the theory of learned non-use, Taub and colleagues observed and described the impact of the increased use of the ipsilesional upper limb in the unilaterally deafferented monkeys [e.g. (Taub, 1976a) (Taub, 1976b; Taub, 1980)]. While bilateral deafferentation led to the continuing use of the affected upper limbs, unilaterally deafferented monkeys had to be forced over a longer period of time in order to persistently reduce compensatory behavior. These observations are corroborated by later studies in rodents [e.g. (Jones and Schallert, 1994; Kerr et al., 2016; Balbinot and Schuch, 2018)] and monkeys [e.g. (Friel and Nudo, 1998; Kaeser et al., 2010; Murata et al., 2015)] employing cortical lesion models giving rise to some possible explanations for the underlying neural mechanisms [reviewed in (Jones, 2017; Buetefisch, 2015)]. We therefore decided to include the aspect of compensation as an addition to the final definition to allow for a comprehensive characterization of the phenomenon.

The synthesis of the information related to the diagnostic process reflected the expected lack of a standardized clinical procedure to identify and differentiate the LNU phenomenon from other sensorimotor or neuropsychological phenomena. The participating experts did however agree on key elements required to optimally gather information building up to the identification of non-use in patients. Following the participants’ responses, the ideal diagnostic approach involves a multidisciplinary team of health professionals in addition to the patient and his/her relatives or caregivers in a longitudinal approach starting in the acute phase. Controversially discussed were the degree of objectivity of information sources available to identify the non-use behavior. While the contrast of the spontaneous against the forced usage was mentioned by mostly all participants, the actual modalities of testing were diversely described and miscellaneous measures and readouts were mentioned. In a small number of answers, concerns were raised as to whether LNU could be diagnosed and unequivocally differentiated from other related clinical pictures such as dyspraxia or hemineglect. It is necessary to point out that attempts to objectify LNU in patient populations have been made and used for experimental purposes (Johnson et al., 2011; Han et al., 2013; Bailey et al., 2015), so far however, these rather elaborate approaches have not been implemented into clinical routine.

Obviously, LNU is still more of a so-called anecdotal clinical observation and not contained in the ICD-10 systematic. Nonetheless, it is necessary to keep in mind, that therapeutic approaches like CIMT are originally legitimized with the non-use phenomenon. However, evidence is accumulating that CIMT, selected here as one example for current therapeutic approaches, is not effective in promoting recovery as a ‘one fits all’ solution for every patient (Nijland et al., 2011; Corbetta et al., 2015). Hence, in order to select the optimal therapeutic strategy for a patient, more knowledge is needed with respect to (a) the identification of the underlying mechanisms that bring about a certain clinical picture of sensorimotor dysfunction, (b) the subsequent stratification of the patient population, and (c) the response rate of a certain therapeutic approach in the presence of a selected sensorimotor dysfunction. These three points call for a wide range of basic and applied research in the field of rehabilitation.

Strikingly, the described diagnostic procedures currently in use lack a systematic verification of the experience-dependent characteristic of the LNU phenomenon, which constitutes one fundamental element of the newly developed definition. While learning mechanisms have been convincingly shown to contribute to non-use in animal models, this important proof is still lacking in human populations. First experimental work using immobilization paradigms in neurologically healthy human volunteers underscore as well as contradict (Ngomo et al., 2012; Rosenkranz et al., 2014; Ikeda et al., 2019) the seminal work of Taub and colleagues. It is obvious that showing the gap between spontaneous and forced use only supports the assumption of a phenomenon of behavioral nature; it does however, not allow to draw conclusions about the mechanisms that regulate its development.

## Conclusion

In conclusion, we emphasize the need for the development of a structured diagnostic procedure and its implementation in the clinical practice, which allows the identification of patients showing or at risk to develop a non-use phenomenon. This step will improve clinical decision making and consequently lead to an optimized therapeutic intervention targeted at the functional recovery of the upper extremity following a brain lesion.

This step may also allow the transfer of elements of the diagnostic procedure to other clinical areas such as chronic pain or complex regional pain syndrome in which LNU like phenomena have been described (Punt et al., 2013).

## Data Availability

The data that support the findings of this study are available from the corresponding author on reasonable request.

## Acknowledgements

We would like to thank all participants of the Delphi study for their commitment to our project, their time, and their motivation to share their knowledge and expertise. This work was supported by the Research Foundation Flanders (FWO) grant (1509816N) to KFH.

## Author contributions

KFH conceived the research. TH, KFH, and MB developed the questions for the Delphi study throughout the whole study. TH implemented and executed the data collection and initial structuring of the data material. TH, KFH, and MB analyzed the data. KFH wrote the first draft of the manuscript, and KFH, TH, and MB revised the manuscript together. All other co-authors contributed with sharing their knowledge and experience in the Delphi study.

